# Forecasting local surges in COVID-19 hospitalizations through adaptive decision tree classifiers

**DOI:** 10.1101/2024.09.12.24313570

**Authors:** Rachel E. Murray-Watson, Alyssa Bilinski, Reza Yaesoubi

**Affiliations:** Department of Health Policy and Management, Yale School of Public Health, New Haven, Connecticut, USA; Public Health Modeling Unit, Yale School of Public Health, New Haven, Connecticut, USA; Department of Health Services, Policy and Practice, Brown University School of Public Health, Providence, Rhode Island, USA

**Keywords:** COVID-19, decision tree classifier, risk prediction, surveillance

## Abstract

During the COVID-19 pandemic, many communities across the US experienced surges in hospitalizations, which strained the local hospital capacity and affected the overall quality of care. Even when effective vaccines became available, many communities remained at high risk of surges in COVID-19-related hospitalizations due to waning immunity, low uptake of booster vaccinations, and the continual emergence of new variations of SARS-CoV-2. Some risk metrics, such as the CDC’s Community Levels, were developed to predict the impact of COVID-19 on the community-level healthcare system based on routine surveillance data. However, they had limited utility as they were not routinely updated based on accumulating data and were not directly linked to specific outcomes, such as surges in COVID-19 hospitalizations beyond local capacities. Regression models could resolve these limitations, but they have limited interpretability and do not convey the reasoning behind their predictions. In this paper, we evaluated decision tree classifiers that were developed in “real-time” to predict surges in local hospitalizations due to COVID-19 between July 2020 and November 2022. These classifiers would have provided visually intuitive and interpretable decision rules for local decision-makers to understand and act upon, and by being updated weekly, would have responded to changes in the epidemic. We showed that these classifiers exhibit reasonable predictive ability with the area under the receiver operating characteristic curve (auROC) > 80%. These classifiers maintained their performance temporally (i.e, over the duration of the pandemic) and spatially (i.e., across US counties). We also showed that these classifiers outperformed the CDC’s Community Levels for predicting high hospital occupancy.

**Significance Statement:** A major concern during the COVID-19 pandemic was the risk of exceeding local healthcare capacity due to COVID-19-related hospitalizations. To assess this risk and inform mitigating strategies, several risk assessment tools were developed during the pandemic. Many of these tools, however, did not predict local outcomes, were not updated as the pandemic progressed, and/or were not interpretable by decision-makers. We propose an adaptive framework of decision tree classifiers to predict whether COVID-19-related hospital occupancy would exceed a given capacity threshold. This framework would provide interpretable classification rules to predict surges in local hospitalizations,and maintained its performance over time and across US counties, and outperformed the CDC’s Community Level tool.

## 1 Introduction

When COVID-19 emerged in the United States in 2020, it proved an immediate threat to healthcare systems. Rapid surges in hospitalizations threatened to overwhelm the healthcare capacity and compromise the standard of care and patient health outcomes [1]. Even after vaccinations were introduced in late 2020, many communities remained at risk of intermittent surges in COVID-19 hospitalizations due to low rates of vaccination, waning infection- and vaccine-induced immunity, and seasonal changes in transmission dynamics [2, 3].

To assess the risk of surges in hospitalizations, significant efforts ware made to predict the trajectory of the pandemic [4], including the COVID-19 Forecast Hub [5], the Scenario Modeling Hub [6], and the IHME COVID-19 Forecast Model [7]. However, the majority of these predictions are made available at the state or national levels [4], reducing their utility in more local settings, where the pandemic trajectory could bear little resemblance to that of the nation or state. Hence, there was a need among local policymakers for tools that will convert the data collected by surveillance systems into meaningful predictions for their area.

One attempt at such a tool was the CDC’s COVID-19 Community Levels [8] (replaced with the COVID-19 Hospital Admission Rate [9] and the COVID-19 County Check [10] in 2023), which was designed to indicate when there may be an upcoming strain on healthcare systems. The COVID-19 Community Levels were based on the number of new weekly COVID-19 cases, weekly hospital admissions due to COVID-19, and weekly inpatient beds occupied by COVID-19 patients and predicted whether the Community Level would be low, medium, or high. While these metrics were chosen for their correlation with future high mortality and hospital occupancy [8], they did not directly predict any specific outcome of interest [11]. Additionally, the thresholds for the Community Levels were chosen once and not updated as the pandemic progressed or new coronavirus strains became dominant. This limited the overall utility of the Community Levels tool in the long term. Previous work has attempted to address these issues by using regression models that are continuously updated to predict concrete outcomes (e.g., mortality level) [12]. The main limitation of regression models is that they are not easily interpretable and do not convey the reasoning behind their assessment of risk for some adverse outcomes (e.g., surges in COVID-19 hospitalizations).

In this paper, we investigate whether accurate decision tree classifiers could be developed to predict local surges in COVID-19 hospitalizations. Decision tree classifiers are machine learning models that provide simple and interpretable classification rules to make predictions [13, 14]. They resemble the threshold-based, flowchart-like structure of the CDC Community Levels, which makes them easy to use and interpret by local policymakers. Using decision tree classifiers, however, allows us to explicitly link the surveillance data to the outcome of interest, which here is to predict whether local hospital occupancy due to COVID-19 patients would exceed a set threshold. To develop these decision tree classifiers, we use data collected from US county surveillance systems and evaluate their performance during different waves of the pandemic between July 2020, before which data was not routinely collected from each county in the US, and November 2022. We also compare the performance of these decision tree classifiers with that of CDC Community Levels.

## 2 Method

### 2.1 Overview

From July 2020, several COVID-19 indicators (e.g., deaths, cases, hospital admissions) were collected through surveillance systems to monitor and predict trends in the pandemic [15, 16]. Our goal is to use these indicators and metrics derived from these indicators as features in decision tree classifiers to predict whether the local hospital capacity is expected to exceed in the short term due to surges in COVID-19 hospitalizations.

We consider four groups of classifiers that differ based on the predictors (features) they use to predict whether local hospital capacity would be surpassed in a three weeks from the current week. These models are described in detail below. To develop and evaluate these models, we used data collected between July 15th, 2020, and November 7th, 2022, a 123-week period. Before July 2020, data was not routinely collected and reported. In December 2022, there was a change in hospitalization reporting guidelines, and data was reported to the Centers for Disease Control and Prevention’s (CDC) National Health Safety Network (NHSN) rather than to the Department of Health and Human Services (HHS). After this period, there were changes in the quality of the data being reported. Hence, we focused our analysis over the period July 15th, 2020 to November 7th, 2022.

For a given week *t*, we train decision tree classifiers using data collected between week 1 through week *t* − 1. We then use the data collected in week *t* to predict the outcome in week *t* + 3. For example, at the beginning of week *t* = 10, we use the data collected through week 9 to develop models to predict whether hospital capacity would be exceeded in week 13. To evaluate how the performance of these classifiers changes throughout the pandemic (especially during the phases where novel variants emerge), we repeat this procedure for every week *t* ∈ {2, 3, 4…, 118}.

### 2.2 Data

We obtained COVID-19 hospital admissions, occupancy, and ICU occupancy data from the Department of Health and Human Services [16], and the data on cases and deaths from the New York Times [15]. Only cases confirmed by a reverse-transcriptase polymerase chain reaction (RT-PCR) test were included in the definition of “case”. We followed the procedures outlined in previous studies for data pre-processing, including the aggregation of weekly observations and the imputation of missing values [11, 12].

To account for patients leaving their county of residence to access healthcare, we aggregated data by Health Services Areas (HSAs)[17] consistent with the CDC’s Community Level calculations. We compiled data at the midpoint of each week. A total of 804 HSAs were included in the analysis; for each of the classifiers we developed, < 1.5% of the health service areas were omitted from the data set due to missing data.

### 2.3 Outcomes

The outcome of interest was whether the COVID-19-caused hospital occupancy would exceed 15 per 100,000 population in exactly three weeks’ time. The capacity threshold of 15 per 100,000 population is calculated in prior studies and falls in the middle of the CDC Community Level’s “Medium” risk assessment for the hospital admissions indicator [18, 19, 20] (Figure S1 in SI). We chose the three-week period for consistency with the CDC Community Levels [8, 12]. However, we note that knowledge of hospital capacity in interceding weeks is also of use to policymakers. Consequently, we considered whether this outcome occurs any week during the interval [*t* + 1, *t* + 3] as a sensitivity analysis. This time period skips the immediate next week but maintains a relatively short period between the training and target weeks.

In addition to the initial threshold of 15 cases per 100,000 population over a three-week period, we performed a sensitivity analysis using alternative threshold values and outcome periods.

### 2.4 Features

The prediction models we consider utilize all or a subset of the following features:

1. Number of COVID-19 cases per 100,000 population in the last week,
2. Number of COVID-19 deaths per 100,000 population in the last week,
3. Number of COVID-19-related hospital admissions per 100,000 population in the last week,
4. Number of hospital beds occupied by COVID-19 patients per 100,000 population in the last week,
5. Number number of ICU beds occupied by COVID-19 patients per 100,000 population in the last week,
6. Portion of hospital beds occupied by COVID-19 patients in the last week,
7. Change in each of the aforementioned metrics in the last week, and
8. Whether the hospital occupancy by COVID-19 patients exceeded the selected capacity threshold in the last week.

### 2.5 Decision tree classifiers

We consider four broad categories of decision tree classifiers, which differ in the features they include, to predict whether hospital capacity will be exceeded:

1. “Naive” classifier that uses only a binary variable of whether the current hospital capacity threshold is exceeded;
2. “CDC Optimized” classifier that uses the same features included in the CDC Community Levels (i.e., new weekly COVID-19 cases, hospital admissions, and the percent of inpatient beds used by COVID-19 patients);
3. “Reduced” classifier, which uses features related to hospital admissions, hospital and ICU occupancy, but not cases or deaths (because as of mid-2023, the county-level case and death data was no longer routinely reported [15]);
4. “Full” classifier that uses all features listed in the previous section.

We compare the performance of these classifiers with that of the CDC’s Community Levels (Figure S1 of SI), which were developed using all data collected between March 1st, 2021 and January 24th, 2022 [8]. To measure the performance of Community Levels, we evaluate it on the period between March 3rd, 2022 and November 20th, 2022, during which its use was encouraged by the CDC.

We also note that the Community Levels designate an area as “low”, “medium”, or “high” risk; hence, to evaluate its ability to predict surges in COVID-19 hospitalizations, we counted all weeks in which the set hospitalization threshold was exceeded as equivalent to a “high” community level, and weeks where it was below as equivalent to “medium/low”.

### 2.6 Model development

For each week *t* between July 15th, 2020 and November 7th, 2022, we use the data collected from all HSAs from week 1 to week *t* − 1 as a single training set to develop our classification trees. We used 10-fold spatial and temporal cross-validation both to optimize the model hyperparameters (see §S2 in SI for details). To ensure that resulting decision trees were easy to interpret, we restricted the depth of trees to less than five layers and then found optimal depth through hyperparameter tuning.

Since the data from larger populations are expected to be less noisy compared to data from smaller populations, we included instance weights in the model fitting procedure, based on HSA population. Additionally, to account for class imbalance in the outcome, we trained the model using “balanced” class weights, with higher weights assigned to the minority class in the decision tree classifier’s function [21, 22].

Though we used 123 weeks of data, due to the 3-week prediction task, the need for one week of test data, and one week for initial training, we could only train 117 decision tree classifiers.

We developed our decision tree classifiers using the scikit-learn package in Python [23].

### 2.7 Model evaluation

In line with Transparent Reporting of a multivariable prediction model for Individual Prognosis or Diagnosis (TRIPOD) recommendations [24], model validation was carried out using temporal validation, partitioning our dataset into training and testing subsets based on time. To evaluate the performance of a decision tree classifier, we calculate the area under the receiver operating curve (auROC) based on data from all HSAs collected during the projection period. A classifier that provide 100% correct predictions has an auROC of 1 and a classifier that randomly guess that outcomes has an auROC of 0.5.

To investigate the regional variation in the performance of our classifiers, we also present the auROC scores based on the out-of-sample predictions by HSA. These auROC scores were calculated based on the predictions made by each weekly classifier for a given HSA, which was then matched with the corresponding HSA.

The auROC metric is agnostic to the selected classification threshold and presume that true/false positives and true/false negatives are equally desirable outcomes. In reality, a predicted surge in hospital capacity will cause different responses among policymakers, each with an associated cost. Under this assumption, the value of true and false predictions may differ. To allow for any differences in preferences policymakers may have, we additionally calculate the net benefit [25], a metric that allows for differences in the weighting of true and false positives, facilitated by an “exchange” parameter, *ω*. The net benefit function incorporating true positive and false positive rates is given by:

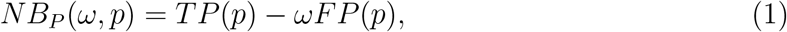

where *TP* (*p*) is the true positive rate and *FP* (*p*) is the false positive rate when the classification threshold *p* is selected (a classifier with classification threshold *p* predicts the hospital capacity would be exceeded if the estimated the probability of exceeding capacity is greater than *p*). A true positive result means that the model has correctly predicted the surge in hospital capacity; hence, any action undertaken by the policymaker to avoid such a surge was justified and could have prevented loss in population health. In contrast, a false positive result could waste resources if it leads to using mitigating strategies. Equation 1 allows policymakers to weigh the economic costs of unnecessary action (*ωFP* (*p*)) with the health benefits of justified action (*TP* (*p*)). These outcomes, however, depend on the classification threshold *p*. To find the optimal classification threshold for a given *ω*, we solve:

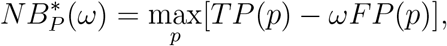

using a grid search over values of *p*.

The above definition of net benefit does not consider the health and economic consequences of true and false negatives. True negatives mean that policymakers can avoid implementing costly interventions and false negatives may risk overwhelming hospital capacity. To account for these factors, we also evaluate our classifiers using the following version of net benefit:

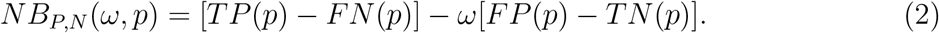

For a given *ω*, we use a grid search to find the optimal classification threshold that maximizes the above function.

We also evaluate the performance of different classifiers using maximum regret (MR). MR is the difference between the performance of the best-performing model and that of the model under consideration and gives a metric of loss or gain in difference in performance by using one model over another. Mathematically, for a given metric *q* (e.g., auROC), the MR of model *m* is defined as [12]:

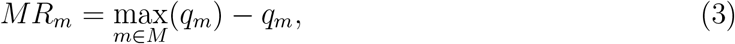

where *M* is the set of all models under consideration and *q*_*m*_ is the performance (e.g., auROC) of model *m*.

Finally, we used SHAP (Shapley Additive exPLanations) values to evaluate the contribution of each feature to predictions [26]. SHAP values provide a way to allocate the contribution of each feature to a model’s prediction by considering all possible combinations of feature values across instances [27]. It provides insight into changes in model performance that may occur if some features are not included. We use the shap package in Python to calculate the SHAP values and present the SHAP summary plot [27]. In this plot, we present the SHAP values for each prediction period across all 117 Reduced classifiers.

## 3 Results

Between July 15th, 2020 and November 7th, 2022, there was a substantial variation in the burden of COVID-19 across HSAs (Figure 1). Throughout this 123-week period, on average, 78% of HSA-weeks surpassed the 15 COVID-19 patients per 100,000 hospitalized threshold (dashed curve in Figure 1C).

**Figure 1.**
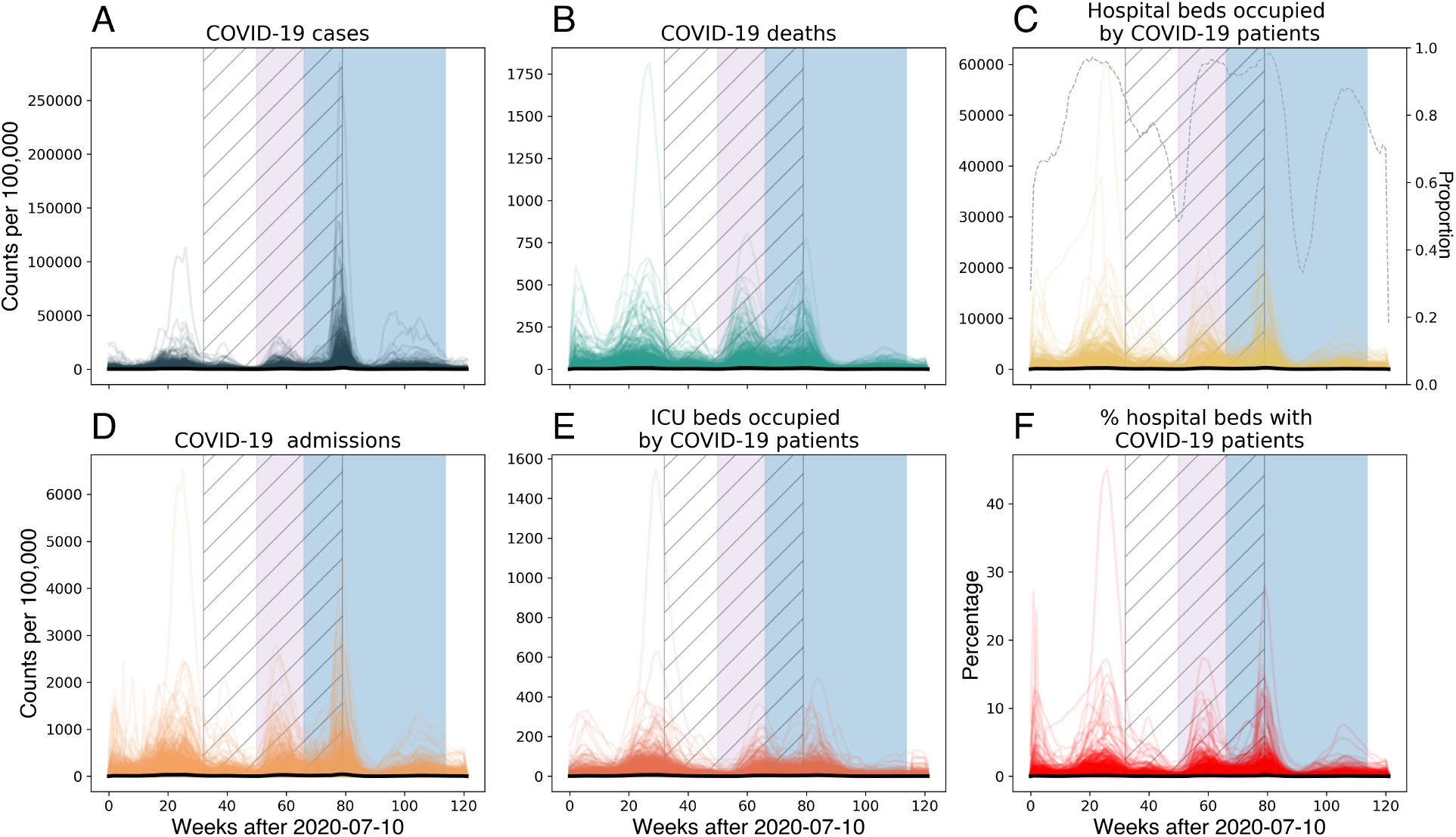
Weekly COVID-19 indicators between July 15th, 2020 and November 7th, 2022 reported by HSAs. The purple and blue boxes shows the period when the delta and omicron variants were dominant, respectively. The hatching shows the data used to develop the CDC Community Levels. The black curve shows the mean across all HSAs for the given indicator.

COVID-19 cases, new hospital admissions, hospital beds and ICU beds occupied by COVID-19 patients, and the percentage of hospital beds occupied by COVID-19 patients were positively correlated with surpassing the hospitalization threshold (Table 1).

**Table 1.** Mean and standard deviation of weekly COVID-19 observations stratified by whether the hospital occupancy is surpassed in 3 weeks’ time or not.

| Feature | Surpassed |  | Not surpassed |  |
| --- | --- | --- | --- | --- |
|  | Mean | Std Dev | Mean | Std Dev |
| Cases | 143.63 | 290.4 | 53.4 | 122.9 |
| Deaths | 1.8 | 3.2 | 1.4 | 3.0 |
| Hospital beds | 37.9 | 87.3 | 1.4 | 4.3 |
| Admissions | 9.4 | 21.6 | 2.3 | 9.1 |
| ICU beds | 1.75 | 3.5 | 0.08 | 0.44 |
| Percent beds occupied by COVID-19 patients | 4.3 | 9.4 | 0.023 | 0.073 |
| Change in cases | -1.4 | 169.0 | 2.5 | 88.1 |
| Change in deaths | 0.059 | 1.1 | -0.16 | 1.3 |
| Change in hospital beds | -0.098 | 41.6 | 0.016 | 4.3 |
| Change in admissions | -0.19 | 14.7 | 0.36 | 9.1 |
| Change in ICU beds | -0.0057 | 1.64 | 0.0076 | 0.38 |
| Change in percent beds occupied by COVID-19 patients | -0.00022 | 0.071 | 0.000024 | 0.0064 |

Across all classifiers, the auROC was always above 0.5, including for the Naive classifiers which used just one feature. The Full classifiers generally had the highest auROC, though the Reduced classifiers were competitive despite not using features related to cases or deaths. The CDC Optimized, Reduced and Full classifiers had an auROC of > 0.8 across all weeks (Figure 2A). The performance of these classifiers was relatively consistent across different waves of the pandemic. However, the regret for auROC scores was best controlled by the Full and Reduced classifiers (Figure 2B).

**Figure 2.**
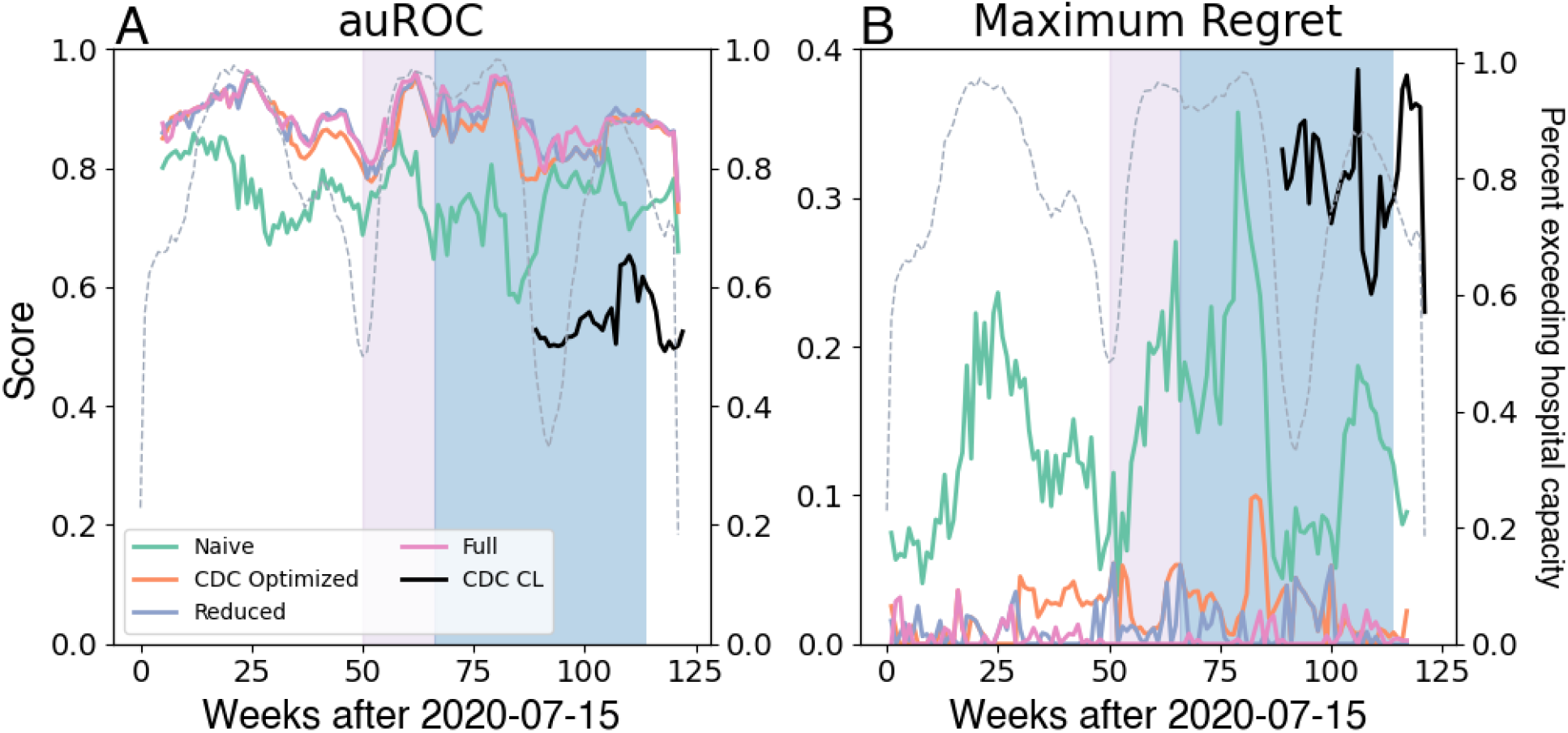
Performance of decision tree classifiers to predict whether the COVID-19 hospital occupancy is expected to exceed 15 per 100,000 in exactly 3 weeks. (A) auROC, (B) maximum regret. The purple and blue boxes show the period when, respectively, the delta and omicron variants were dominant. The gray dashed line shows the proportion of HSAs that exceed the hospitalization threshold of 15 per 100,000 population for a given week.

Across all models, around week 62, there was a decrease in the auROC score, which coincided with when the omicron strain became the predominant strain in the US. There is additionally a decrease in performance when there was a peak in cases during the omicron wave (around week 90). Conversely, the highest auROC scores were achieved when a high proportion of HSAs exceeded the 15 per 100,000 hospitalization threshold (Figure 2A). As county-level case and death data ceased being available from mid-2023, we focused the remainder of the analysis on the Reduced classifiers.

For 76% of the counties, the auROC that could be calculated using the Reduced classifiers over the entire study period exceeded 0.80 (Figure 3). However, for around 23% of counties, an auROC could not be calculated as across each 117 outcome weeks, the hospital beds occupied by COVID-19 patients always exceeded the 15 per 100,000 threshold, or never did. Thus, there was no “true negative” with which to calculate the auROC.

**Figure 3.**
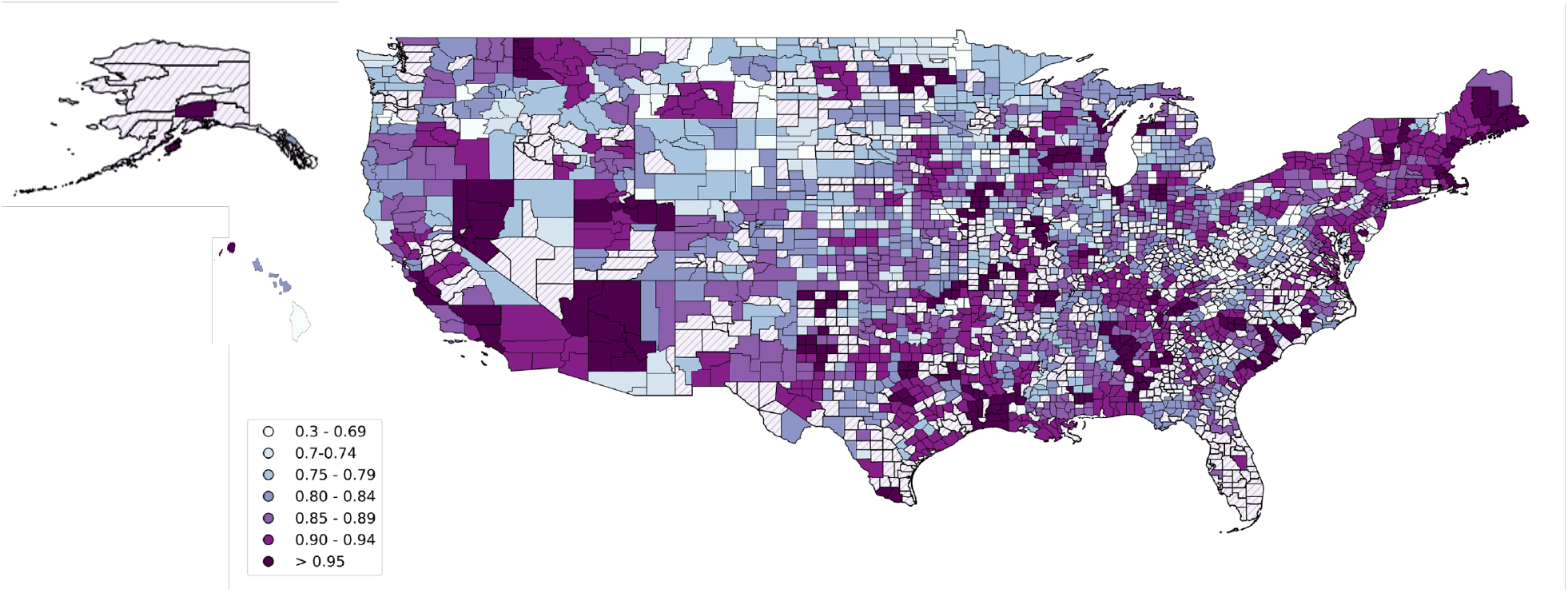
Performance of Reduced classifiers across all counties. The auROC was calculated by HSA using the predictions from all 117 Reduced classifiers. The hatching indicates where there were no true negative instances with which to calculate the auROC, and the auROC is recorded as NA. See Figure S7 for the spatial performance of classifiers that includes case and death data.

The added benefit of the Reduced classifiers compared with the Naive classifiers varied over the pandemic weeks and depended on the selected penalty value (*ω* in Eq. 1) to model the trade-off between the true positive and false positive rates (Figure 4A). For smaller values of *ω*, which represent scenarios where the false positive is minimally penalized, the added benefit of using the Reduced classifiers is unimportant. Reduced classifiers provide greater benefit when false positives are penalized more substantially and when the proportion of HSAs where hospital capacity exceeds 15 per 100,000 is large (Figure 4A). However, the Naive classifiers provide additional net benefit over the Reduced classifiers when the proportion of HSAs exceeding the hospital occupancy threshold was low. The Full classifier presented higher net benefit compared to the Naive classier over each penalty value and weekly classifier (FigureS8 in the SI).

**Figure 4.**
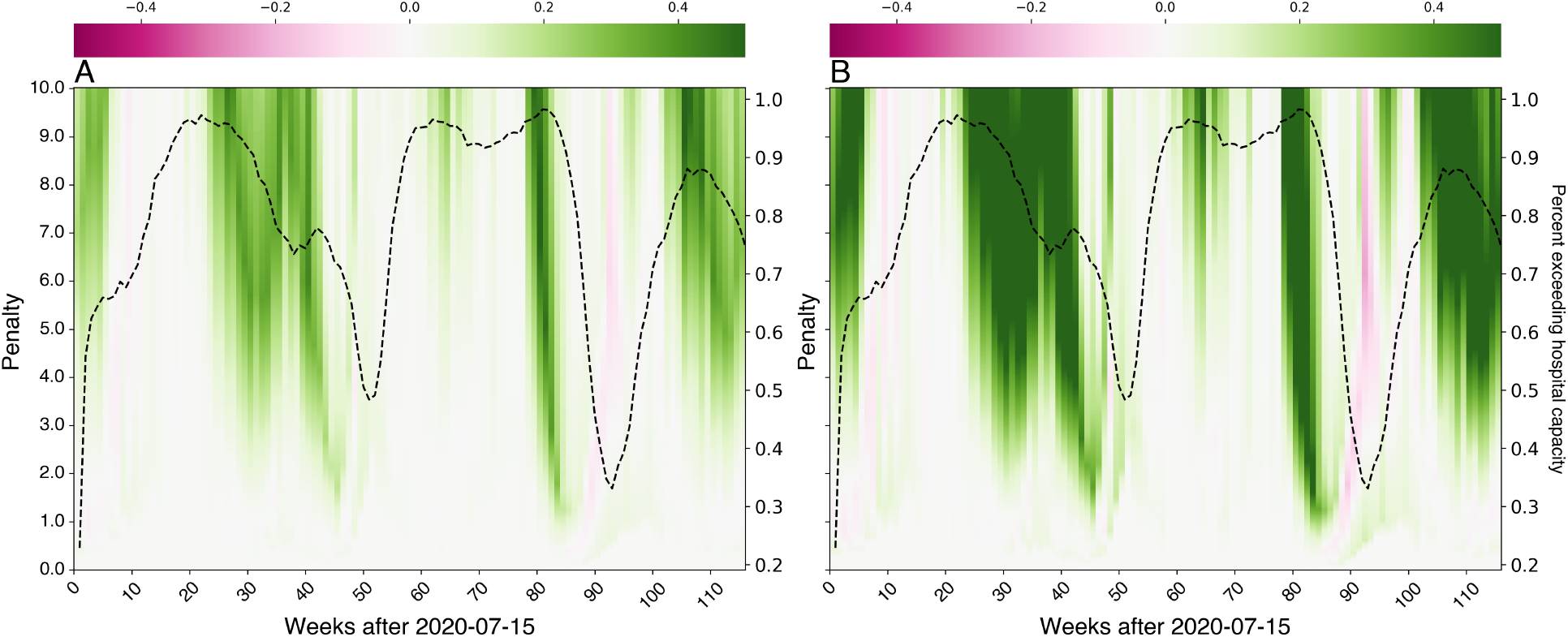
The net benefit of the Reduced classifiers related to the Naive classifiers. (A) Using the net benefit function *NB*_*P*_(), which accounts for false positive and true positive rates and (B) Using the net benefit function *NB*_*P,N*_ (), which accounts for true and false positive rates and true and false negative rates. In areas shaded green, the Reduced classifiers outperform the Naive classifier, while areas shaded pink indicate where the Naive classifier perform better. The gray dashed line is the proportion of HSAs that exceed the 15 per 100,000 hospital capacity for a given week.

We observe similar behavior when the net benefit function *NB*_*P,N*_ (), which accounts for true and false positive rates and true and false negative rates, is used (Figure 4B). When this net benefit function is used, the Reduced classifiers outperform the Naive classifiers for a larger number of weeks and penalty values.

The change in the number of COVID-19 hospital admissions in the previous week, the number of beds occupied by COVID-19 patients, and the number of COVID-19 hospital admissions, were selected as important features in more than 50% of 117 Reduced classifiers between July 15th, 2020 and November 7th, 2022 (Figure 5). Whether the current hospital capacity exceeded the 15 per 100,000 threshold was not included as an important feature in any classifier, despite the good performance of the Naive classifier, which only used this feature. However, this binary feature is highly correlated with other features such as the number of hospital beds by week and the percentage of beds occupied by COVID-19 patients (Table 1), which do appear as important features (Figure 5).

**Figure 5.**
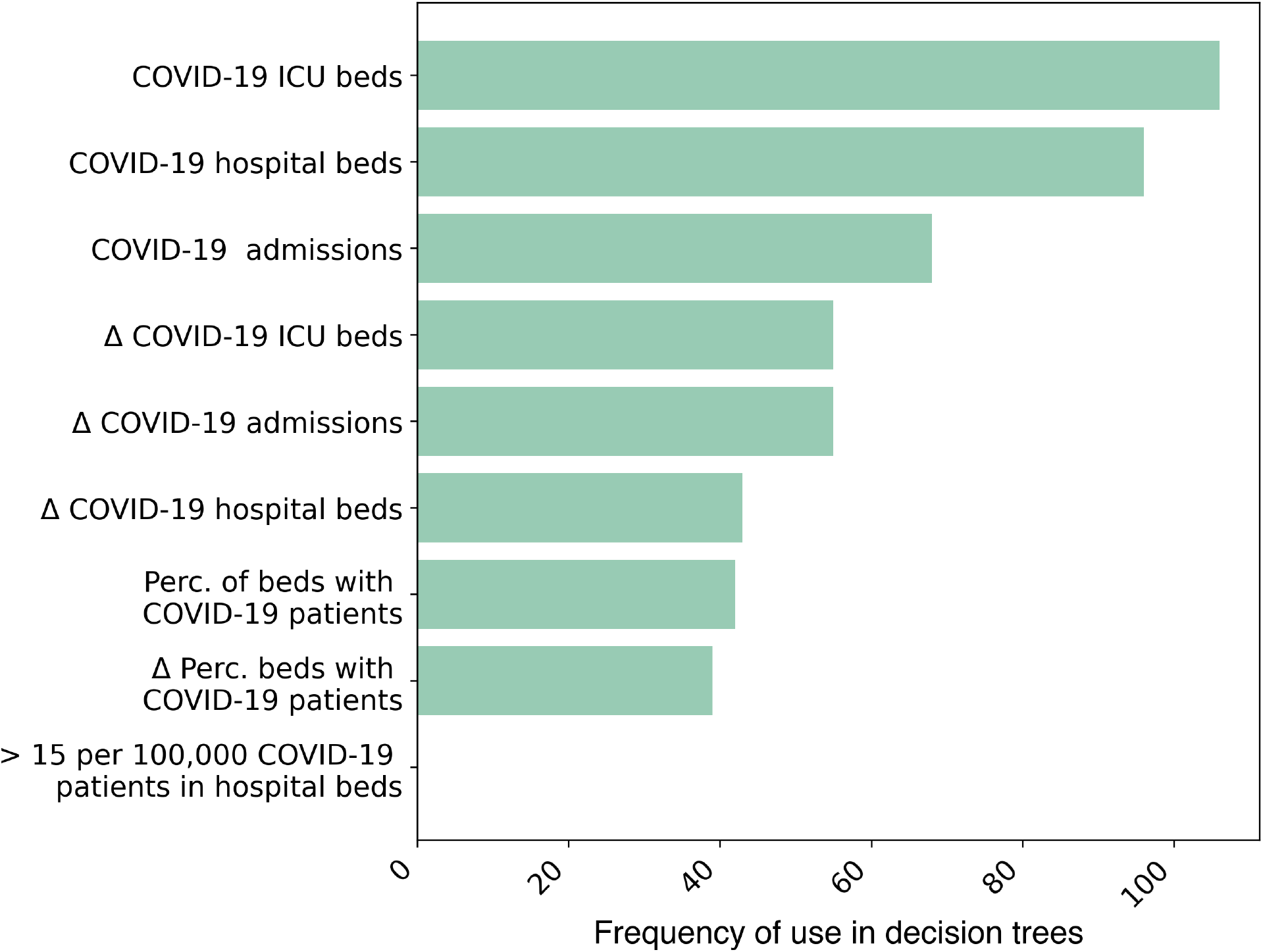
Frequency at which a feature was identified as important in 117 Reduced classifiers created between July 15th, 2020 and November 7th, 2022.

The SHAP values also indicate that COVID-19 admsissions and ICU beds occupied by COVID-19 patients have a large influence on model predictions (Figure 6). The SHAP values broadly follow a trend where high numbers of admissions and occupied hospital beds (indicated by the pink dots in Figure 6) increase the log odds of a positive prediction, whilst lower admissions decrease the log odds.

**Figure 6.**
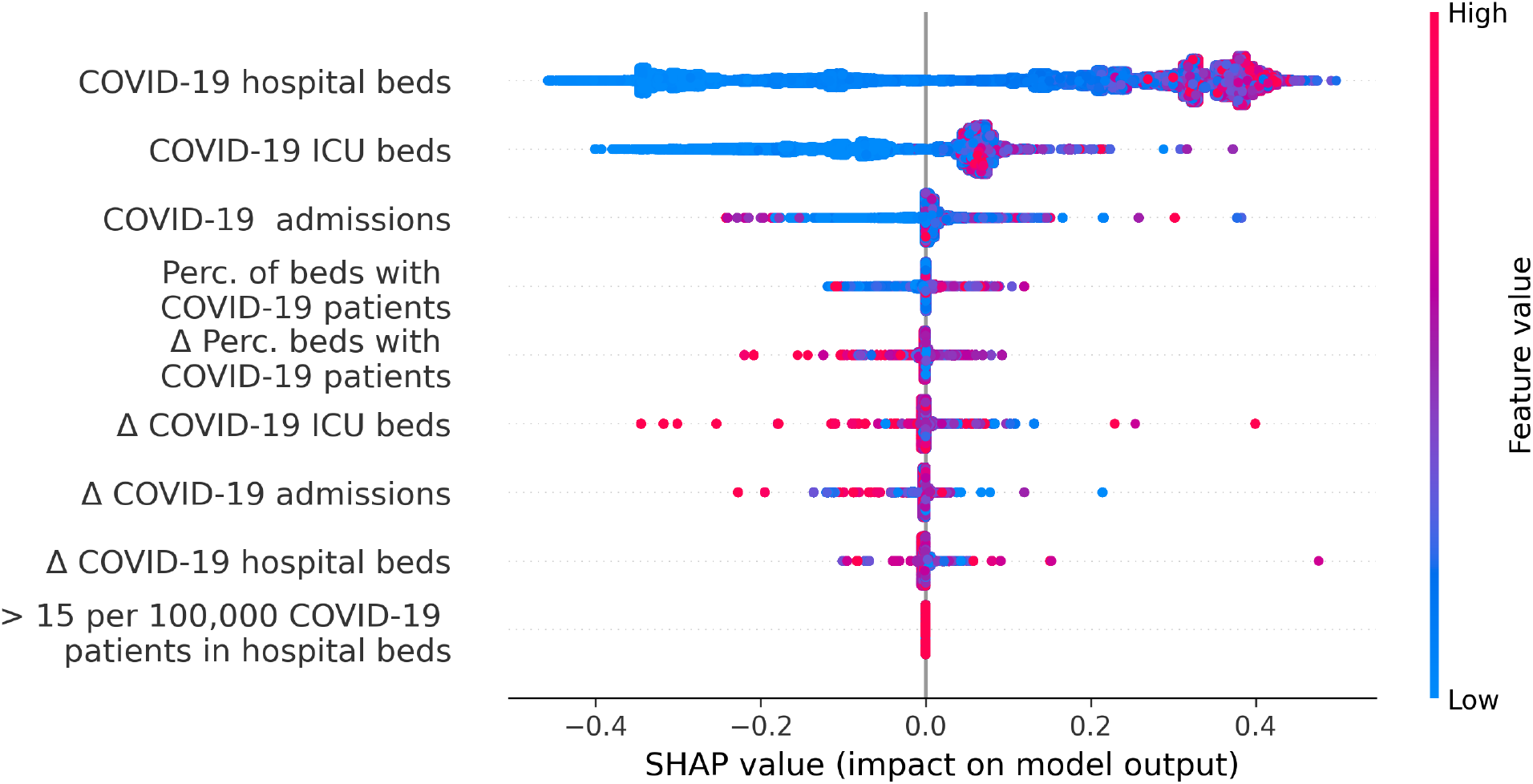
SHAP summary plot. Each point represents the SHAP value for a feature and an instance (observations from a single HSA over a single week). The color of each point represents the value of the feature from low to high. Overlapping points are jittered vertically to display the distribution of SHAP values per feature.

One major advantage of and the main motivation for the use of decision tree classifiers is that they are interpretable. To demonstrate, we present three decision tree classifiers developed for three different stages of the pandemic (Figure 7): the week of July 14th, 2021, when the delta variant was circulating in the population but was not yet the dominant strain (Panel A), the week of August 4th, 2021, when the delta variant was dominant (Panel B), and the final week in our study time period, i.e., the week of November 20, 2022 (Panel C). To illustrate how these decision tree classifiers could be used in practice by a local policymaker, we consider the scenario observed in an HSA in Maryland for July 14th, 2021 (Table 2). Since hospital beds occupied by COVID-19 patients = 22.82, which is less than 29.0, the condition of the first decision node is satisfied. Hence, we check the condition of the second node, whether hospital beds occupied by COVID-19 patients ≤ 10.0, which is satisfied. Therefore, this classification decision tree predicts that the hospital capacity of 15 per 100,000 population is not expected to be exceeded in 3 weeks’ time.

**Table 2.** Data for Allegany/Garrett HSA in Maryland on July 14, 2021. This HSA contains Allegany and Garrett counties in Maryland and Grant, Hardy, and Mineral Counties in West Virginia.

| Value |  |
| --- | --- |
| Hospital Admissions | 1.35 |
| ICU beds | 0.52 |
| Hospital beds | 22.82 |
| Percent beds occupied by COVID-19 patients | 1.5 |
| Change in admissions | -1.81 |
| Change in ICU beds | 0.34 |
| Change in hospital beds | 5.65 |
| Change in percent beds occupied by COVID-19 patients | 0.38 |
| Currently exceed threshold capacity | Yes |
| Exceed capacity three weeks later | Yes |

**Figure 7.**
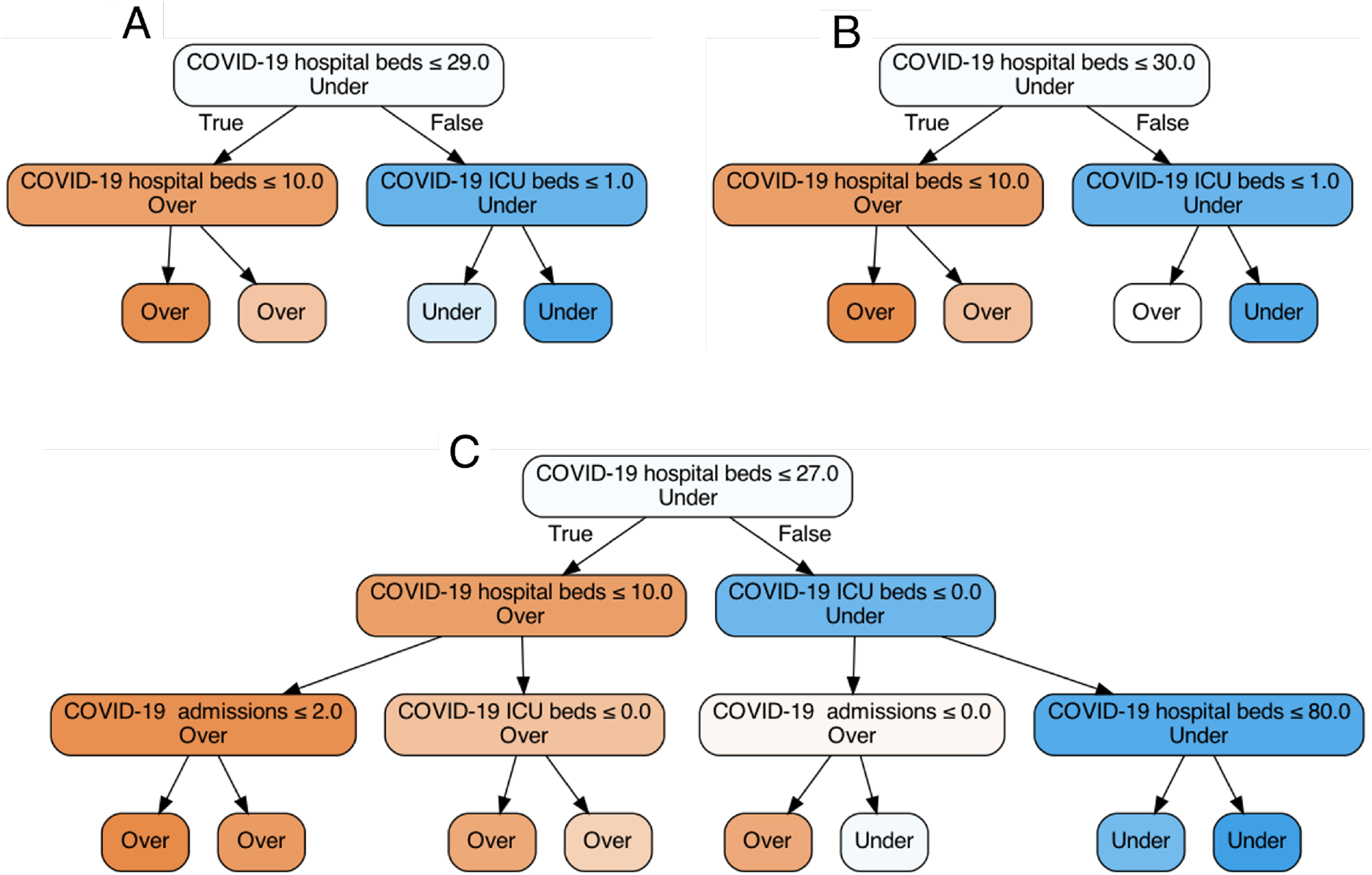
Decision tree classifiers developed at three different points during the pandemic. The week of July 14th, 2021, when the delta variant was circulating in the population but was not yet the dominant strain (Panel A), the week of August 4th, 2021, when the delta variant was dominant (Panel B), and the final week in our study time period, i.e., the week of November 20, 2022 (Panel C).

We note that while these classification trees are developed for different phases of the pandemic, they all use the same features: 1) the number of hospital beds occupied by COVID-19 patients, 2) the number of ICU beds occupied by COVID-19 patients, and 3) the number of hospital admissions of COVID-19 patients. What is different between these classification trees is the classification thresholds and the depth of the tree; for example, the classification threshold at the first node of these trees are 29.0 (Panel A), 30.0 (Panel B), and 27.0 (Panel C).

## 4 Discussion

We presented an adaptive framework to generate simple classification rules that predict whether COVID-19 hospitalizations would have exceeded local capacity. These classification rules are easy to communicate and use the latest data from surveillance systems that are available at the local level. To develop these classification rules during the weeks of July, 2020 to November, 2022, we trained decision tree models on “expanding” datasets, where all available data up to the target week of interest are used in the training procedure. This allowed for the model training procedure to account for changes in pandemic trajectories as population-level factors not directly accounted for as features in the model (e.g., vaccination rates, waning immunity, and easing non-pharmacological interventions). Our analysis suggests that classification rules that are adaptively updated could maintain their predictive ability temporally (i.e., over the duration of the pandemic; Figure 2) and spatially (i.e., across US counties, Figure 3). Additionally, these classifiers outperform other classifiers trained only on more recent datasets (see SI S4).

Our classifiers were trained on a range of COVID-19 indicators that were routinely reported between July 2020 and November 2022, including hospital admissions, and hospital occupancy. However, the source of case and death data used in this study for the “Full” classifiers ceased being updated in mid-2023 [15], and other data sources are only updated at the state level [28] if at all [29, 30]. Though this limits the training data on which the classifiers can be trained, we showed omitting death and case data does not meaningfully impact the performance of the characterized classification rules (Figure 2). Indeed, the CDC Optimized, Full, and Reduced presented reasonable predicted accuracy, with an auROC of > 0.80 for the majority of classifiers; even classifiers based solely on current occupancy can achieve relatively high auROC, including when a new strain becomes dominant in the population (Figure 2). In the Reduced classifiers, the admissions and change in admissions, and occupied ICU beds were most frequently considered to be important in the decision tree classifiers (Figure 5). Similarly, these features had some of the largest explanatory effects on the auROC (Figure 6).

We focused on providing predictions at the level of HSAs as this would have enabled local policymakers to detect potential surges in COVID-19 hospitalizations and to respond accordingly before the local hospital capacity is overwhelmed. One such previous attempt was the CDC’s Community Levels framework. This framework, however, lacked a direct relationship with a specific outcome of interest [18] and was never updated after its development. Other studies have provided more systematic classification rules predicting specific outcomes (such as hospital capacity [20] or high mortality [12]). We extended this work by demonstrating that decision tree classifiers trained on surveillance data could predict whether COVID-19 hospitalizations may exceed the local capacity level with reasonable accuracy. These classification rules provided by these models are easy to use and interpret in practice (Figure S9 in SI).

The spread of SARS-CoV-2 varied substantially across different communities and geographic regions, depending on the implementation of non-pharmaceutical interventions [31], local rates of vaccination [32], and the emergence of novel strains, amongst other factors [33, 34, 35]. Despite the substantial heterogeneity in data reported by HSAs, our analysis suggests that our Reduced classifiers maintained their performance across US counties (Figure 3). We note that the spatial performance of classifiers was not dramatically improved when case and death data were used to develop classification rules (Figure S7).

By using the net benefit framework, our approach allows for incorporating policymakers’ preferences between different prediction outcomes (i.e. false negative and positive and true negative and positive). For example, predicting that there will be a surge in hospital occupancy when there will not actually be one (false positive) or predicting no surge when one is going to occur (false negative) have distinct health and economic costs; the former could have high economic costs and the latter could lead to high adverse health outcomes. Our analysis shows that the classification rules identified by our approach present positive net benefit values, especially during the phases of the pandemic when the COVID-19 hospitalizations were rising or declining (Figure 4).

Two of the classifiers considered here (i.e., “CDC Optimized” and “Full”) use case and death data. The available case data undercounted the actual number of infections, as only cases confirmed by a molecular laboratory test were included [15]. Limited testing availability [36], particularly at the beginning of the pandemic, and asymptomatic transmission contributed to this undercount [37]. Similarly, death counts relied on the patient to be diagnosed according to guidelines specified by state and federal bodies [38], which could have led to both the mis- and underdiagnosis of patients. However, our analysis shows that even without these COVID-19 indicators (as in our “Reduced” classifier), reasonable predictive power can still be achieved (Figure 2).

An important limitation of predictive models that are only trained on historical data is that the historical data may not sufficiently capture the possible changes in pandemic trajectories due to future events. This is particularly true if novel strains emerge with characteristics that are substantially different from what is manifested in historical data. The use of simulated trajectories, which can be parameterized to allow for such variations, may help to make the predictions of the classifier more robust [20].

The proposed approach is flexible to facilitate a number of extensions. While we trained our models to predict a binary outcome, classification decision trees could also be developed to predict multiple outcomes, such as “low”, “medium” and “high” levels of COVID-19 hospital capacity, if thresholds to define these outcomes are available. Furthermore, we only included indicators related to the COVID-19 pandemic to predict surges in hospital occupancy. However, there is evidence of cocirculation of influenza, COVID-19, and respiratory syncytial virus infections during recent winters [39, 40], which threatens to overwhelm the healthcare system. Data from surveillance systems related to these infectious diseases could be incorporated to provide more robust predictions. Finally, while we focused on predicting surges in hospitalizations, other metrics of interest, such as ICU capacity or mortality, could also be considered depending on the availability and the quality of data related to these outcomes.

## Data Availability

All data produced in the present work are contained in the manuscript and available online at https://github.com/nytimes/covid-19-data and https://healthdata.gov/Hospital/COVID-19-Reported-Patient-Impact-and-Hospital-Capa/anag-cw7u/about_data

