## Supplemental Material for "Forecasting local surges in COVID-19 hospitalizations through adaptive decision tree classifiers"

### Supplementary information

#### S2 Hyperparameter tuning and cross-validation

To avoid over-fitting and to ensure that the classifiers are robust to temporal and spatial variation in our dataset, we used the following cross-validation approach to tune hyperparameters of each classifier for each week $t$:

(1) Divide the dataset by HSAs into ten validation sets, denoted by $F_1, F_2, \dots, F_{10}$:

(2) For each validation set $F_i, i \in \{1, 2, \dots, 10\}$:

  (i) Do a 10-fold cross-validation to optimize hyperparameters using the remaining dataset (excluding $F_i$). Let $h_i^*$ denote the optimized hyperparameters (Table **??**) for this fold.

  (ii) Evaluate the performance (e.g., auROC) of the optimized model on the validation set $F_i$. Let $p_i^*$ denote this estimated performance.

(3) Return the set of hyperparameters with the highest $h_i^*$.

The ranges for the hyperparameters are shown in Table S1.

| **Parameter** | **Range** |
|---|---|
| criterion | gini, entropy |
| Max depth of tree | 2 - 5 |

Table S1: Parameter Grid for Decision Tree

#### S3 CDC Community Levels

| **New Cases** (per 100,000 population in the last 7 days) | **Indicators** | **Low** | **Medium** | **High** |
|---|---|---|---|---|
| **Fewer than 200** | New COVID-19 admissions per 100,000 population (7-day total) | <10.0 | 10.0-19.9 | ≥20.0 |
| | Percent of staffed inpatient beds occupied by COVID-19 patients (7-day average) | <10.0% | 10.0-14.9% | ≥15.0% |
| **200 or more** | New COVID-19 admissions per 100,000 population (7-day total) | NA | <10.0 | ≥10.0 |
| | Percent of staffed inpatient beds occupied by COVID-19 patients (7-day average) | NA | <10.0% | ≥10.0% |

**The COVID-19 community level is determined by the higher of the inpatient beds and new admissions indicators, based on the current level of new cases per 100,000 population in the past 7 days**

Figure S1: **Criteria for establishing CDC Community Levels** [1].

#### S3.1 Evaluating classifiers on entire CDC Community Level training period

In the main text, we evaluate the performance the CDC Community Levels, on the data collected between March 3rd, 2022, and November 25th, 2022. These are compared with the performance of our decision tree classifiers that are trained on all data collected between week 1 and week $t-1$. To allow for a more fair comparison, we trained our decision tree classifiers on the same data as the original Community Levels, collected between March 1st, 2021, to January 24th, 2022. The first two-thirds of the data were used to train the classifiers, and they were evaluated on the remaining third.

Though 95.8% of the weeks designated a "high" Community Level did exceed the hospital capacity threshold, 87.2% of weeks predicted to be "medium" and 71.0% of the "low" risk weeks also did. When we designated the weeks predicted to be "high" and where the hospital capacity exceeded the designated threshold as "true positives", the CDC Community Levels have an auROC of 0.663, a value lower than those obtained using the decision tree classifiers above (Figure S 2).

Using the same training and test data, the CDC Optimized classifiers had improved predictive power over the original Community Levels (auROC = 0.818). However, the Reduced classifier had the best predictive power when trained and evaluated during this 47-week period(Table S2). The predictive power of these classifiers was comparable to those of the adaptive classifiers developed on the continually-updated data sets.

Table S2: Comparison of performance of the CDC Community Levels and the decision tree classifiers trained and tested between March 1st, 2021 and January 24th, 2021.

| **Model** | **auROC** | **Maximum Regret (auROC)** |
|---|---|---|
| CDC Community Levels | 0.660 | 0.224 |
| Naive | 0.720 | 0.184 |
| CDC Optimized | 0.817 | 0.087 |
| Reduced | 0.904 | 0 |
| Full classifier | 0.887 | 0.017 |

The CDC Optimized classifier is solely based on the percentage of beds occupied by COVID-19 patients. Where the features overlap with the CDC Community Levels, the thresholds differ (Figure S9).

### S4 Sensitivity Analysis

#### S4.1 Change in prediction task

In the main text, we focused on predicting whether capacity will exceed a given threshold in week $t + 3$. Here we present the evaluation metrics for models trained on a "shifted" time period, where the prediction task is to predict hospital capacity over a three-week period subsequent to $t$ ($[t + 2, t + 5]$). The auROCs for the Full, CDC A, and Reduced classifiers trained to predict this outcome were comparable with the prediction of capacity in week $t + 3$ ( $> 0.80$; Figure 2 in the main text). The auROCs of the Naive classifiers was lower, but still above 0.60 across the 117 weeks.

##### S4.1.1 Change in hospitalization threshold

In addition to the hospital capacity threshold of 15 per 100,000 people that was used to generate our binary outcome in the main text, we also explored two other thresholds: 10 or 20 per 100,000 people. The models were trained in accordance with the procedure outlined in the main text, though now the feature and outcome relating to hospital capacity are replaced with a binary variable calculated based on the new threshold of interest.

In each case, when the Full or Reduced classifier was used, a high auROC was achieved. Generally, the model's performance declined when predicting the lower threshold of 10 per 100,000 (Figure S3), potentially because at most stages during the study period, the majority of HSAs exceeded this threshold. The models were better at predicting the higher threshold of 20 per 100,000. Overall, though performance did vary between thresholds, the overall auROC scores were high, suggesting that the model training procedure is robust to different outcomes of interest.

##### S4.1.2 Change in duration of outcome period

In the main analysis, we predicted whether the hospital capacity would exceed 15 per 100,000 in three weeks' time. We additionally investigated three other periods: two ($t$+2), four ($t$+4), or six ($t$+6) weeks. Shorter outcome periods benefited the performance of the model (Figure S4), particularly when there was a decrease in the proportion of HSAs that exceeded capacity (around week 90).

##### S4.1.3 Limiting the size of the training dataset

The models in the main text have an "expanding" training set, which includes all available data up to the test week, $t$. We tested out more limited training sets, namely restricting the training data to a four-week ($[t - 5,\ t - 1]$), ten-week period ($[t - 11,\ t - 1]$) and a twenty-six-week period ($[t - 27,\ t - 1]$). When we included a larger dataset, it actually had negative impacts on the performance of the model (Figure S5), with the auROC of the classifier trained on twenty-six weeks of data often worse than the equivalent naive four-week model.

##### S4.1.4 Training models every four weeks

As COVID-19 remains a comparatively new disease, and vaccination levels remain heterogeneous and relatively low across the country, we predict hospital capacity over the short term as there remains a risk that the healthcare system will be overwhelmed. We also investigate the frequency at which the decision tree classifiers should be retrained with new, more recent data to ensure their continued accuracy.

Rather than deliver predictions every week, we trained models to deliver predictions every four weeks using all data available up to $t$, and predict over the next four weeks and associated outcome periods. In this scenario, the performance metrics varied less week-to-week (Figure S6), though the classifiers still had reduced performance when there were large changes in the proportion of HSAs where the hospital capacity exceeded 15 per 100,000, though overall performance was comparable to that of the weekly models.

### S5 SI Figures

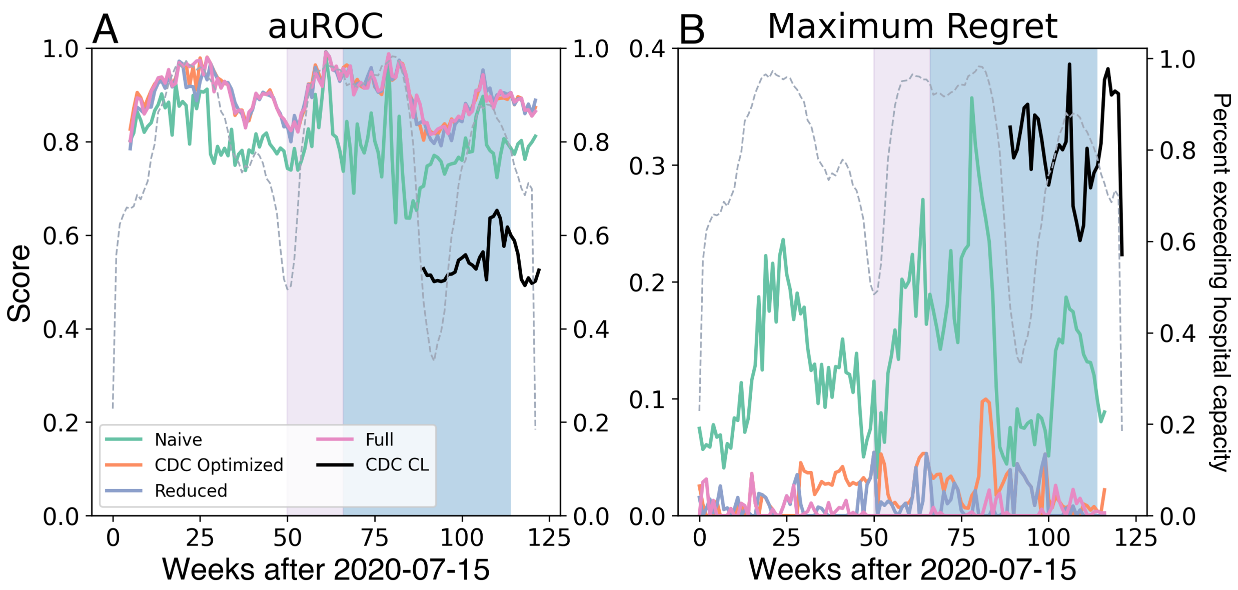


Figure S2: **Performance of decision tree classifiers** when the outcome of interest is whether the hospital capacity exceeds 15 per 100,000 in the “shifted” three-week period. (A) auROC, (B) maximum regret of the auROC. The pink shaded box shows when the delta strain was dominant, the blue when the omicron strain was. The gray dashed line shows the proportion of HSAs that exceed the hospitalization threshold for a given week.

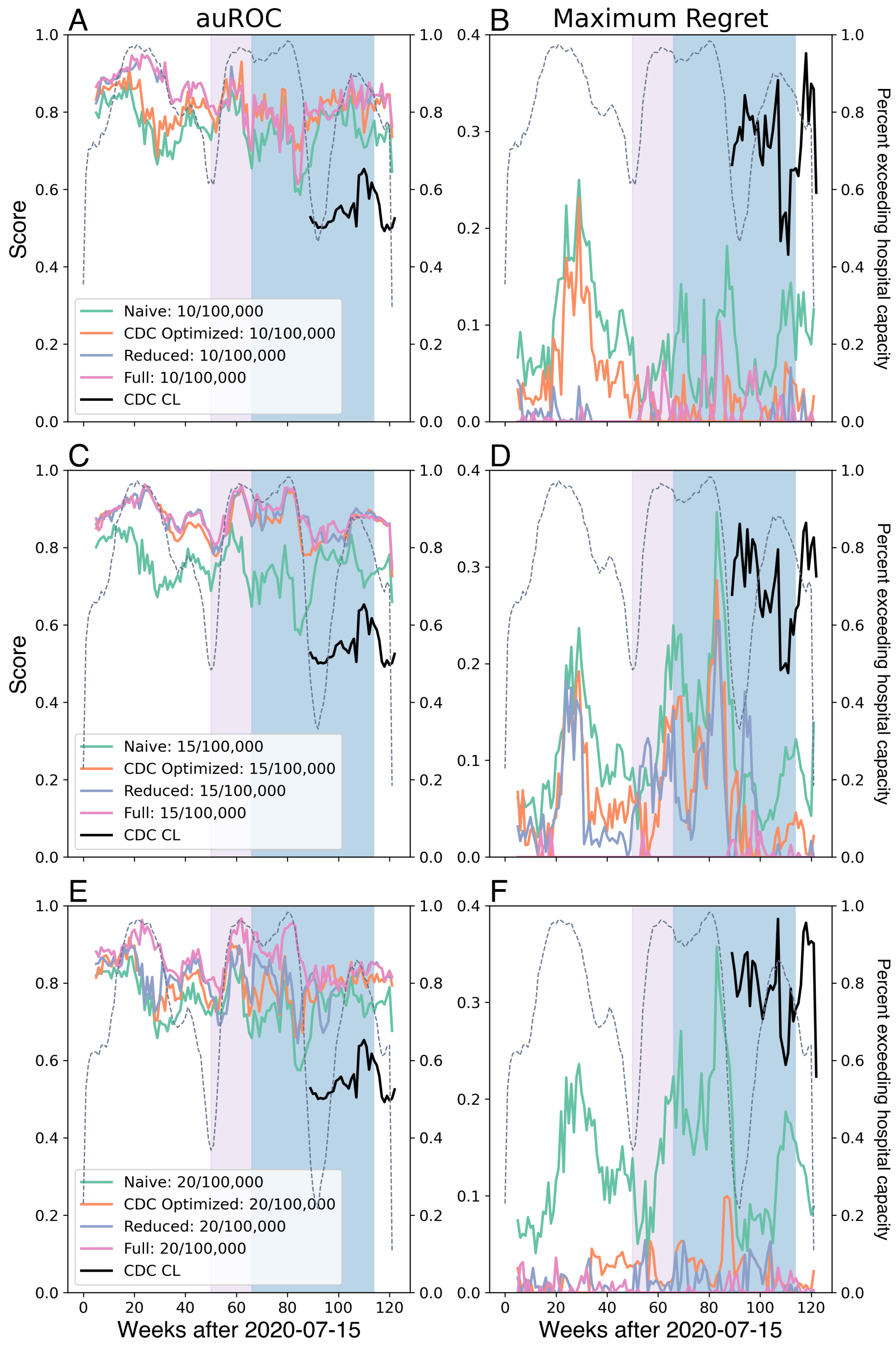


Figure S3: **Performance of decision tree classifiers** when the outcome of interest is whether the hospital capacity exceeds either 10 or 20 per 100,000 during the outcome period. (A) The auROC, (B) the maximum regret. The pink shaded box shows when the delta strain was dominant; the blue when omicron was. The dotted gray line shows the proportion of HSAs that exceed each given hospitalization threshold.

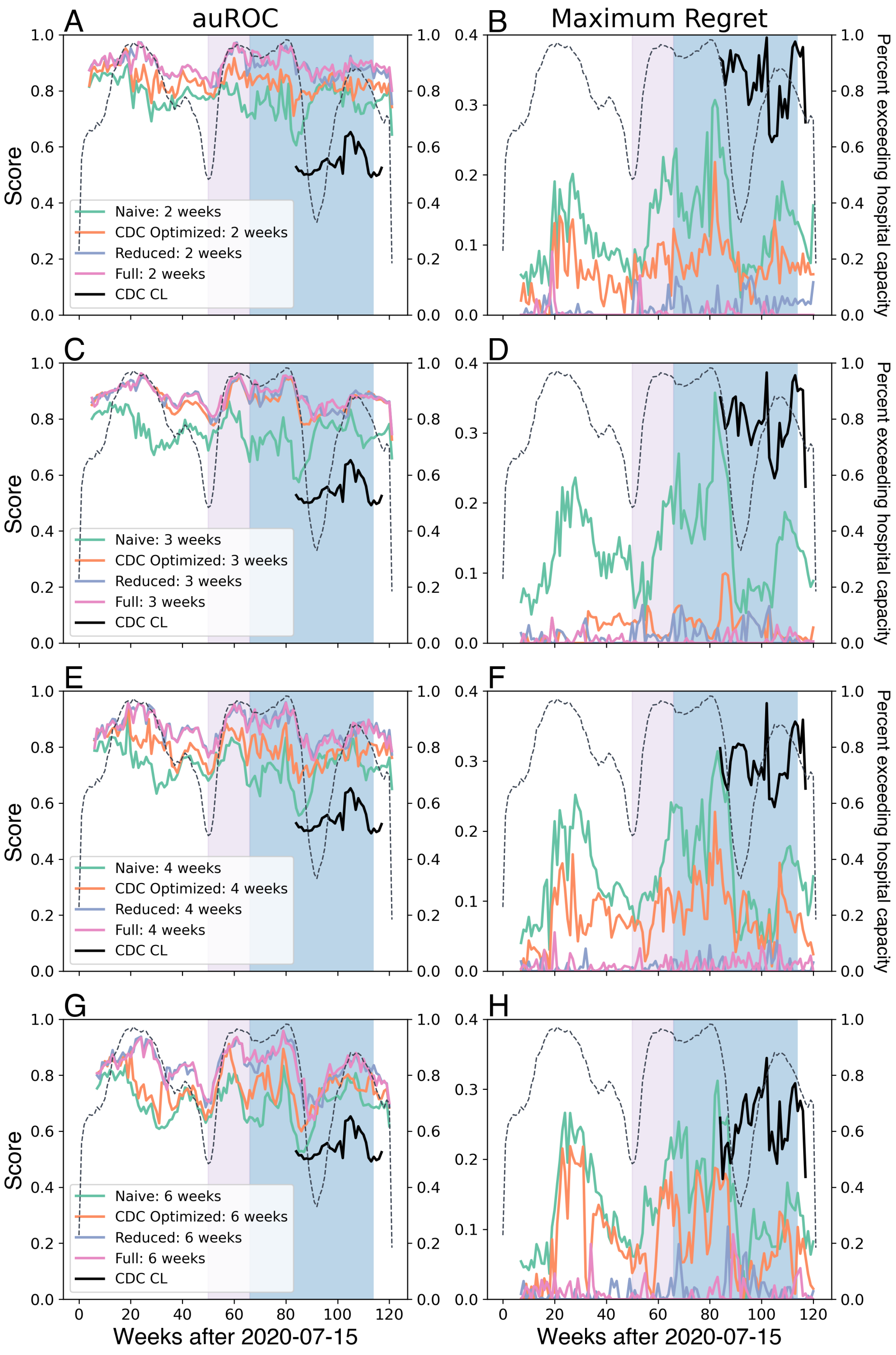


Figure S4: **Performance of full decision tree classifiers** when the outcome of interest is whether the hospital capacity exceeds 15 per 100,000 people in over a 2, 3, 4, or 6 week period. (A), (C), (E), (G) show the auROC, while (B), (D), (F), (H) show the maximum regret per the auROC for each outcome period. These models are developed on an using all data previous to the target week of interest. The pink shaded box shows when the delta strain was dominant; the blue when omicron was. The dotted gray line shows the proportion of HSAs that exceed each given hospitalization threshold.

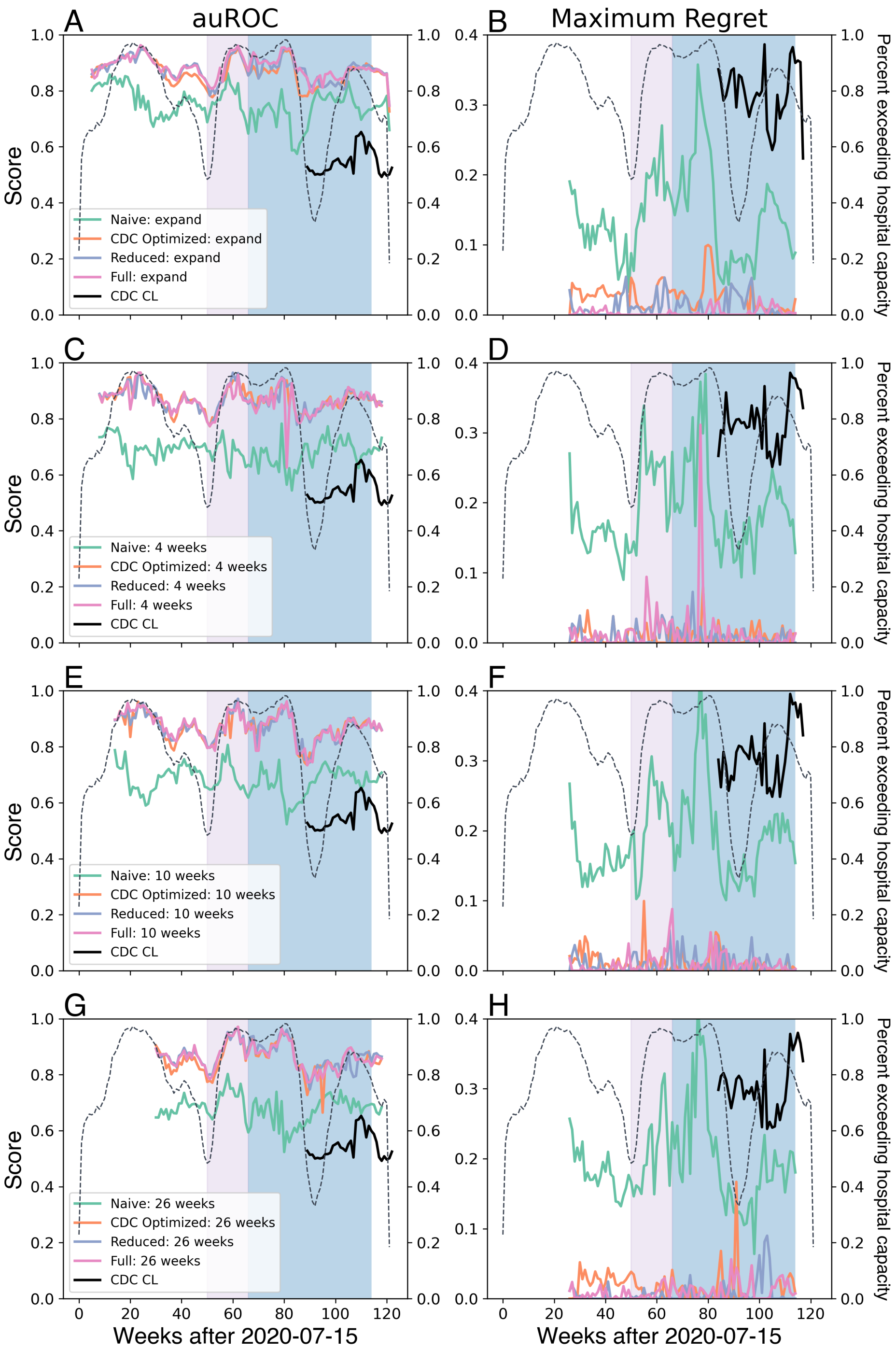


Figure S5: **Performance of full decision tree classifiers** when the training set is either the previous 4, 10, or 26 weeks. (A), (C), (E), (G) show the auROC, while (B), (D), (F), (H) show the maximum regret per the auROC for each training period. The pink shaded box shows when the delta strain was dominant, the blue when the omicron strain was. The dotted gray line shows the proportion of HSAs that exceed each given hospitalization threshold.

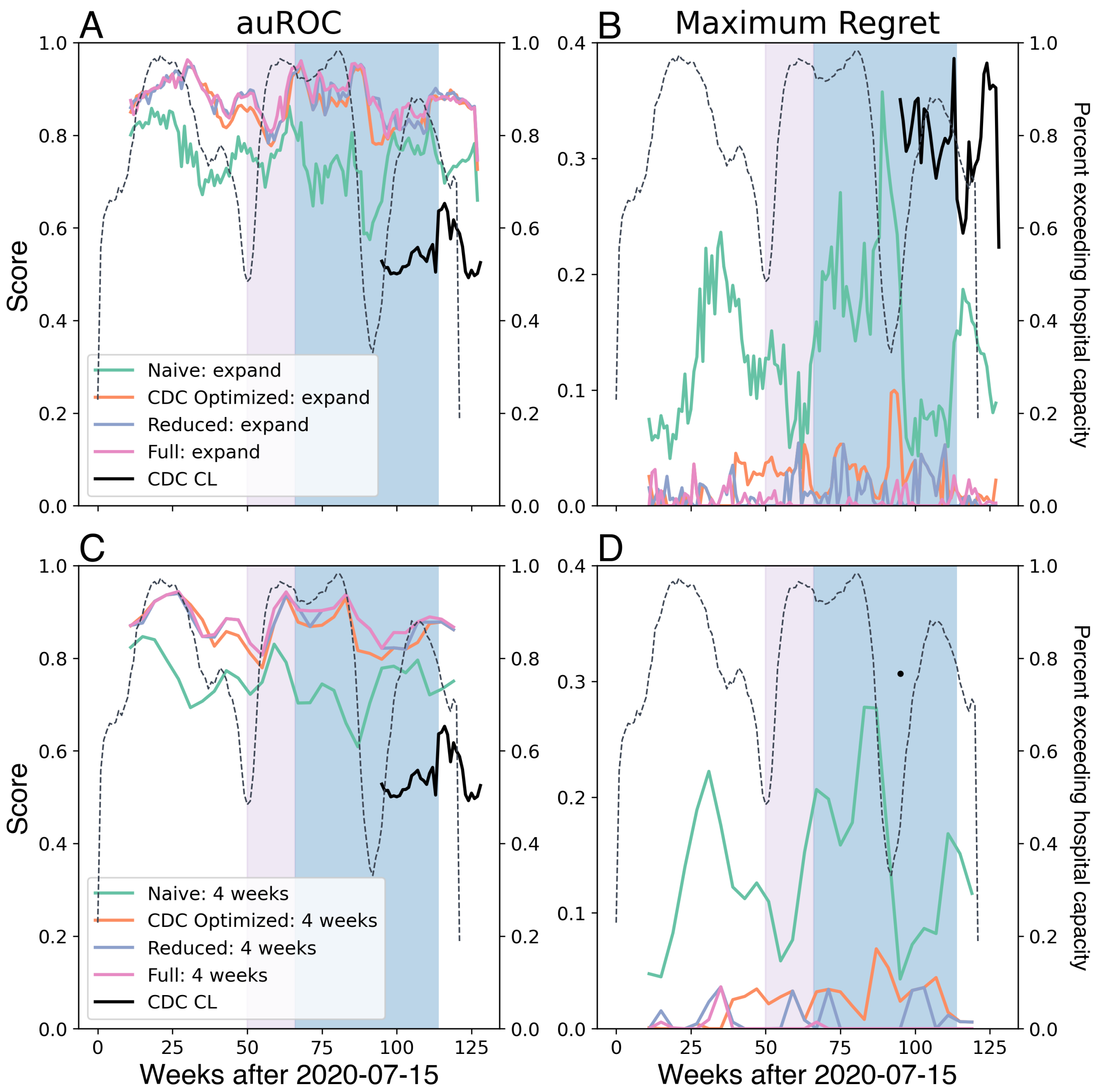


Figure S6: **Performance of decision tree classifiers** when the model training set is only updated every four weeks, and predict whether hospitalizations will exceed capacity over the next four outcome periods. (A) and (C) The auROC, (B) and (D) maximum regret for the auROC. The pink shaded box shows when the delta strain was dominant; the blue when omicron was. The dotted gray line shows the proportion of HSAs that exceed each given hospitalization threshold.

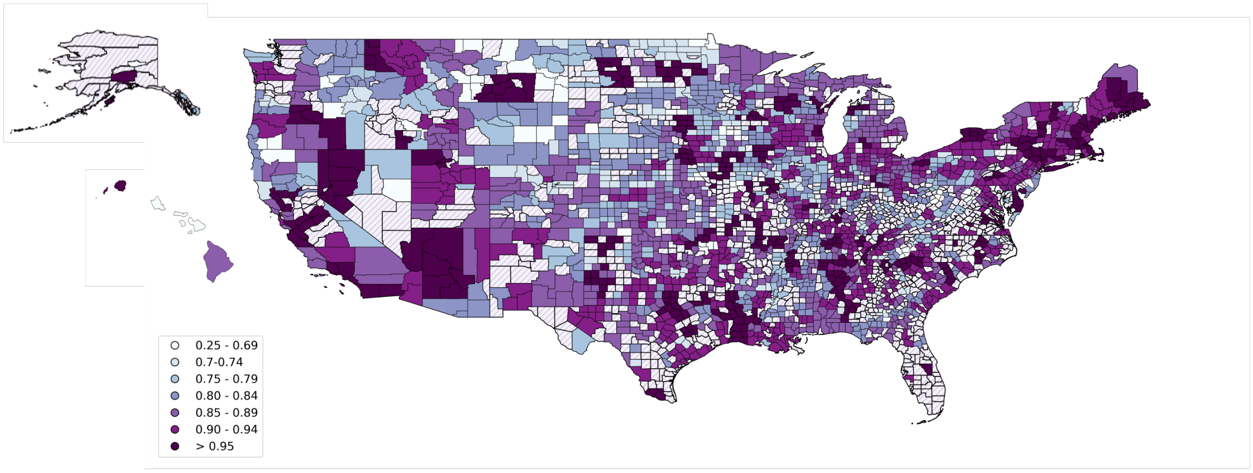


Figure S7: **Performance of the decision tree classifiers across all HSAs when COVID-19 death and case data is used**. The auROC was calculated by HSA using the predictions from all 177 Full classifiers. The hatching indicates where there were no true negative instances with which to calculate the auROC, and the auROC is recorded as NA.

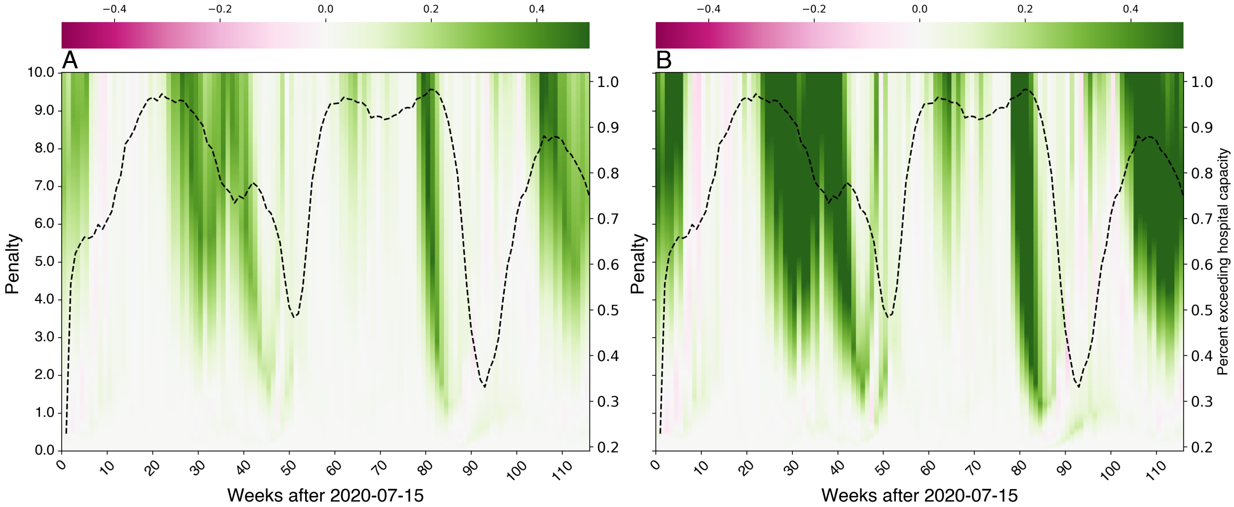


Figure S8: **The net benefit of the Full classifiers related to the Naive classifiers.** (A) Using the net benefit function $NB_P()$, which accounts for false positive and true positive rates and (B) Using the net benefit function $NB_{P,N}()$, which accounts for true and false positive rates and true and false negative rates. In areas shaded green, the Full classifiers outperform the Naive classifier, while areas shaded pink indicate where the Naive classifier performs better. The gray dashed line is the proportion of HSAs that exceed the 15 per 100,000 hospital capacity for a given week.

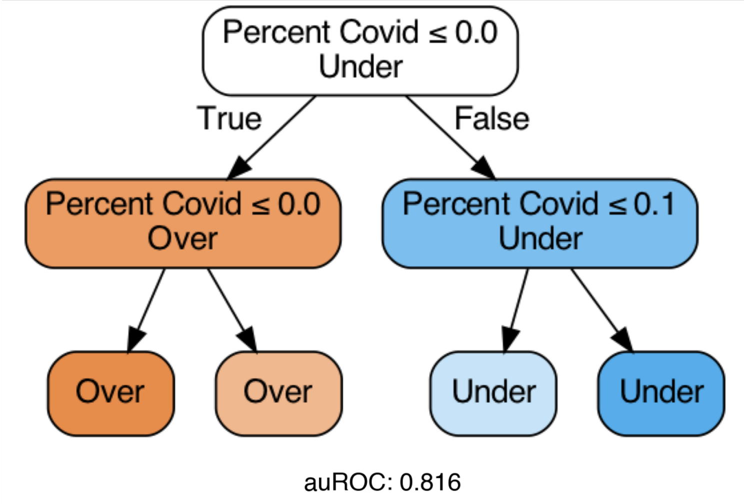


Figure S9: **Decision tree classifiers trained on data from March 1st, 2021 to January 24th, 2021.** (A) Using the same features as the CDC Community Levels.
